# Outcomes of ureteroscopy and internal ureteral stent for pregnancy with urolithiasis: a systematic review and meta-analysis

**DOI:** 10.1101/2020.09.21.20198705

**Authors:** Xingwei Jin, Boke Liu, Yunqi Xiong, Weichao Tu, Yuan Shao, Lin Zhang, Dawei Wang

## Abstract

**Objectives:** To investigate the outcomes of internal ureteral stent versus ureteroscopy (URS) treatments for pregnant women with urolithiasis.

**Design:** This is a systematic review and meta-analysis of observational studies that investigated the outcomes of internal ureteral stent and ureteroscopy for pregnancy with urolithiasis. This systematic review have been registered on the PROSPERO website (www.york.ac.uk/inst/crd, registration number: CRD42020195607).

**Data Sources:** Relevant studies published from January 1980 to April 2020 were identified through a systematic literature search in MEDLINE, EMBASE, Web of Science and the Cochrane Library.

**Data extraction and synthesis:** All pregnant women in their all pregnancy stages who were underwent double-J (D-J) stent insertion only or URS operation for lithotripsy/stone extraction/exploration were considered. The number of related participants in study more than 10 were included. Fertility outcome and complications of intervention were extracted as main outcomes, while other data such as operation success rate, stone free rate (SFR), patient characteristics, anaesthetic method, ureteral stone characteristics, detail of interventions were obtained as well. Complications were stratified according to Clavien-Dindo criteria. Two authors independently extracted data and assessed the quality of included studies. Study-specific prevalence rates were pooled using a random-effects model. We applied the Newcastle-Ottawa Scale quality assessment to evaluate the quality of the selected studies.

**Results:** A total of 25 studies were identified with 131 cases undergoing serial stenting and 789 cases undergoing URS operation. The age range was from 16 to 41, and urolithiasis occurred in the second trimester most. Ultrasound was the most commonly used diagnostic method. The most common site of calculi was distal ureter. The average stone size was between 6-17mm. There were 6 studies investigating D-J stent insertion only, while 23 studies involving URS operation. The most commonly used anesthesia for internal ureteral stent therapy was local anesthesia, and for URS treatments, general anesthesia and spinal anesthesia were widely used. The pooled operation success rate was 97% for D-J stent insertion, and 99% for URS. Serial D-J stenting was an effective methods for treating ureter obstruction and only a few patients passed stone spontaneously. Different common lithotripters were used in URS operations and the pooled SFR was about 91%. For internal ureteral stent therapy; the rate of normal fertility outcome was 99%, but the pooled incidence of complications was about 45%. For the URS treatment group, the rate of normal fertility outcome was 99% as well, and the pooled incidence of complications was about 1%. However, the pooled premature and abortion incidence rate of two group were the same as less than 1%, and the same as this in serious complication incidence rate.

**Conclusions:** Both ureteroscopy operation and internal ureteral stent were usually used for handing pregnancy with urolithiasis. Two treatments had less side effective on fertility outcome, but internal ureteral stent may cause more complications. Evidence suggests that URS therapy may have a greater advantage for pregnancy with urolithiasis when the conditions permit. As it is proved safe and effective, internal ureteral stent could be considered at emergency condition or preoperative preparations was lack.

## INTRODUCTION

The incidence rates of pregnant women with symptomatic urinary tract stones is reported as range from 1 in 2000 to 1 in 200[1]. Symptomatic urolithiasis can lead to renal colic, urinary tract infection and ureteral obstruction posing significant morbidity and potentially mortality not only to mother but also to child. The main risks are pre-term delivery and premature rupture of membranes, which brings serious health risks to the fetus[2, 3]. It is important for the urologists and obstetricians to be aware of the management of this condition.

When managing a pregnant patient with urolithiasis, conservative management is favoured where possible. Surgical intervention are available for those that do not improve with conservative measures [4]. Ureteroscopy (URS) and internal ureteral stent are the most widely used in pregnancy with symptomatic urolithiasis[5]. Insertion of double-J (D-J) stent till definitive treatment in the postpartum period is a temporising measure and related studied is not so many. And with continued advancements in endoscopic technology and endourological techniques, URS seems to be considered as first-line treatment in the management of ureteric stones in pregnancy. However, although the latest 2020 European Association of Urology (EAU) guideline has recommended URS as reasonable alternative option [6], there is still lack of evaluation of evidence-based medicine in comparison between URS and internal ureteral stent. This systematic review and meta-analysis tried to update the outcomes of internal ureteral stent and URS treatments for pregnant women with urolithiasis and make a comparison.

## METHODS

We performed the systematic review according to a predetermined protocol and reported in accordance with the Preferred Reporting Items for Systematic Reviews and Meta-Analyses Protocols (PRISMA-P) guidelines[7]. We had registered our systematic review on the PROSPERO website (www.york.ac.uk/inst/crd, registration number: CRD42020195607). Two reviewers independently undertook the literature search (X.J. and B.L.), assessment for eligibility (X.J. and B.L.), data extraction (Y.S. and W.T.), and qualitative assessment (D.W. and Y.X.). Any inconsistencies between the two reviewers were reviewed by a third reviewer (L.Z.) and resolved by consensus. By consensus among all three reviewers (X.J., B.L. and L.Z.), if data sources were duplicated in more than one study, only the original study was included in the meta-analysis.

### PICOS definition of this study

Participants: Pregnant women with urolithiasis whatever which pregnancy stage they were.

Intervention: D-J stent insertion only.

Comparators (controls): URS operation for lithotripsy/stone extraction/exploration.

Outcome: Fertility results and complications.

Study design: RCTs and observational studies (case-control, cross-sectional and cohort) were included in this systematic review and meta-analysis.

#### Eligibility criteria

1). Pregnant women in any pregnancy stages who underwent D-J stent insertion only or ureteroscopy operation for urolithiasis treatment were included. 2). Studies published between January 1980 and April 2020 were eligible for evaluation. 3). The number of related participants in each group of study should be more than 10.

Studies were excluded if they: 1). Article type including review, comment, letter, guideline, or meta; 2). Related data of pregnancy or interventions was lack; 3). Photographic skill, equipment evaluation or diagnosis criteria of urolithiasis in pregnancy; 4). Research for neonates; 5). Physiologic hydronephrosis without stone disease; 6). Extracorporeal shock wave lithotripsy, percutaneous nephrostomy, or other treatments for pregnancy with urolithiasis.

### Search strategy

We conducted a literature search using PubMed (MEDLINE), Embase, Web of Science and the Cochrane Library which were published from January 1980 to April 2020. The Medical Subject Heading (MeSH) terms were used in conjunction with the following keywords for our search: (Pregnanc∗ or Pregnancy or Pregnant or Gestation∗ or Pregnant woman or Mother∗)**AND** (Urinary Calcul∗ OR Urinary Calculi OR Urinary Calculus OR Urinary Stone∗ OR Urinary Tract Stone∗ OR Ureteral Calcul∗ OR Ureteral Calculi OR Ureteral Calculus OR Kidney Calcul∗ OR Kidney Calculi OR Kidney Calculus OR Nephrolith OR Renal Calcul∗ OR Renal Calculi OR Renal Calculus OR Kidney Stone∗ OR Staghorn Calcul∗ OR Staghorn Calculi OR Staghorn Calculus OR Urinary Lithiasis) **AND** (Ureteroscopies OR Ureteroscopic OR Ureteroscopic Surgical OR Ureteroscopic Surgical Procedure∗ OR Ureteroscopic Surgery OR Ureteroscopy) **AND** (Double-J stent OR Ureteral stent OR Ureteral double-J stent OR Ureteral D-J stent OR Double J ureteral stent OR D-J ureteral stent OR stent OR D-J stent). Full search strings are presented in **Table S1**. References from relevant articles, editorials, conference abstracts, letters, and reviews were thoroughly reviewed to identify additional studies. Full manuscripts of every article with a relevant title and abstract were then reviewed for eligibility.

### Data extraction and qualitative assessment

Two reviewers (Y.S., W.T.) independently extracted the following study-level characteristics from each eligible study: first author, year of publication, country where the study was conducted, journal, study period, age, trimester, diagnose method, stone location and size, anesthetic method, intervention and sample size, operation success rate, stone free rate (SFR), fertility outcome, complications and follow-up pattern. Two groups were set as different treatment procedures: internal ureteral stent (D-J stent) therapy group and URS group.

Fertility outcome and complications were also assessed with Clavien-Dindo classification which as showed in **Table S2**. Clavien-Dindo III-V was regarded as serious complications.

We applied the Newcastle-Ottawa Scale (NOS) quality assessment tool to evaluate the quality of the selected observational studies. This tool was used to measure the key aspects of the methodology in selected studies with regard to design quality and the risk of biased estimates based on three design criteria: 1) selection of study participants; 2) comparability of study groups; and 3) assessment of outcome and exposure with a star system (with a maximum of 9 stars). We judged studies that received a score of 7-9 stars to be at low risk of bias, studies that scored 4-6 stars to be at medium risk, and those that scored 3 or less to be at high risk of bias. A funnel plot was used to assess the publication bias. Any disagreement on the data extraction and quality assessment of the studies were resolved through comprehensive discussion (D.W., Y.X. and L.Z.).

### Statistical analysis

Study-specific prevalence rate estimates were combined using a random-effects model, that considers within-study and between-study variations. Corresponding 95% Confidence Interval (CIs) were extracted directly from articles where available. The statistical heterogeneity among studies was evaluated using Cochran’s Q test and *I*^*2*^ statistic, with values of 25%, 50%, and 75% representing low, moderate, and high heterogeneity, respectively. The criterion for identifying heterogeneity was a *P* value less than 0.05 for the Q test.

An estimation of publication bias was evaluated by the Beggs funnel plot, in which the SE of log (OR) of each study was plotted against its log (OR). An asymmetrical plot suggests possible publication bias. Egger’s linear regression test assessed funnel plot asymmetry, a statistical approach to identify funnel plot asymmetry on the natural logarithm scale of the rates. All statistical analyses were performed using Stata (version 14.2; StataCorp LP, College Station, Texas). All *P* values were two-sided, and *P* <0.05 was considered as statistically significant.

## RESULTS

### Selection of studies

A detailed PRISMA flow diagram of literature search and inclusion criteria were shown in **Figure 1**. A total of 453 studies were initially identified with this literature search (123 from Pubmed, 147 from Embase, 144 from Web of Science and 29 from Cochrane Library).198 studies were excluded due to duplication and 208 were excluded after screening the titles and abstracts. 22 other studies were excluded after full-text review. Finally, a total of 25 studies were identified as eligible for systematic review and meta-analysis.

**Figure 1.**
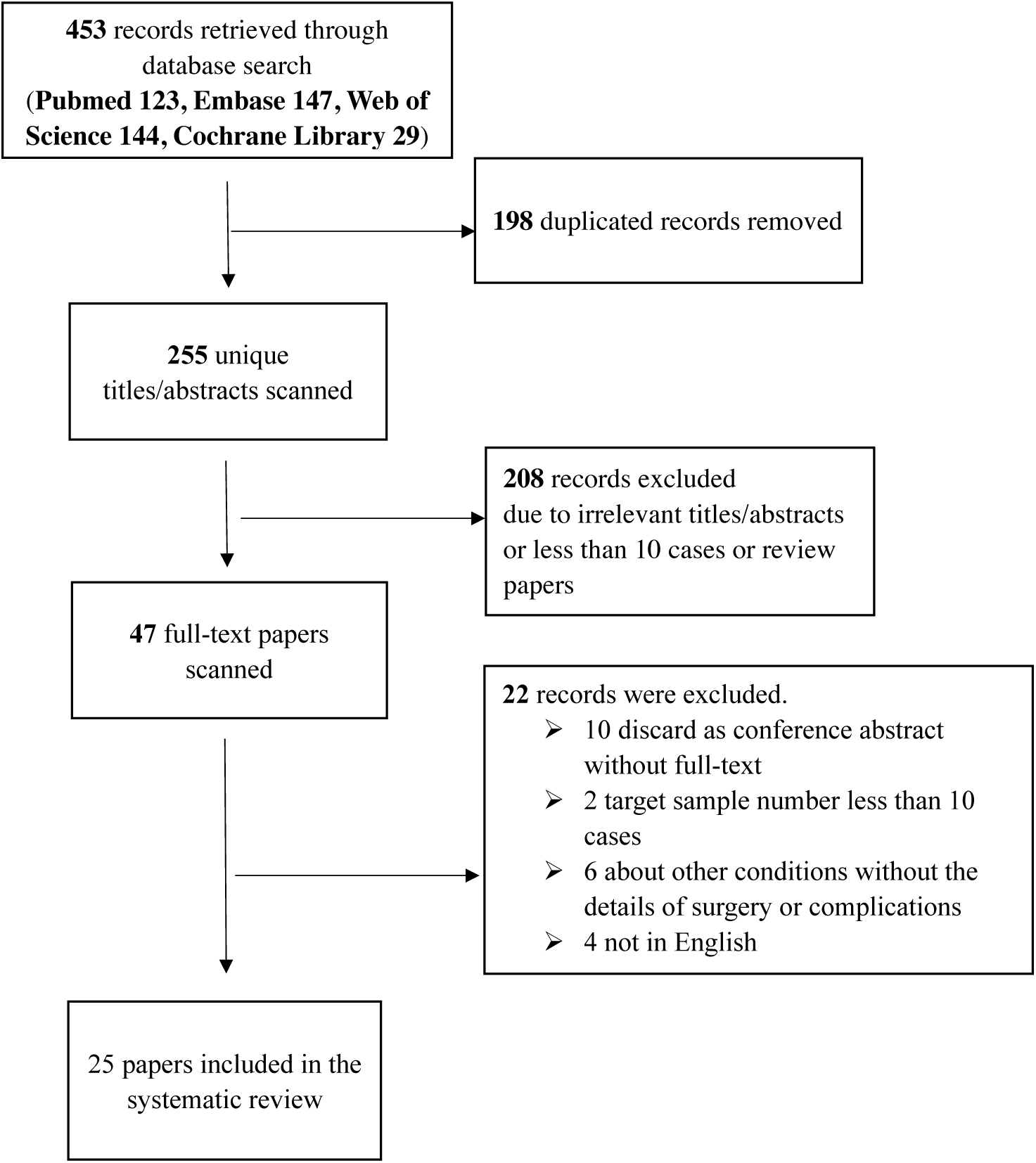
PRISMA flow diagram of study selection for meta-analysis

The published time span of twenty-five studies included was year 1995-2018, and the research period of cases was between 1984 to 2016. Common information of publications was showed in **Table 1**. Briefly, among these studies, 1 from Norway[8], 1 from Italy[9], 2 from America[10], 1 from Brazil[11], 1 from Pakistan[12], 4 from Egypt[13, 20, 27, 29], 5 from China[14, 22, 28, 30, 32], 6 from Turkey[15-18, 21, 25], 2 from Iran[23, 31], 1 from Iraq[24] and 1 from Romania[26]. The age range was from 16 to 41, and urolithiasis occurred in the second trimester most. Ultrasound was the most commonly used diagnostic method. The most common sites of calculi were as follows: distal ureter, medium ureter, proximal ureter. The average stone size was between 6-17mm.

**Table 1.** Summary of characteristic for studies included in the meta-analysis.

| First author | Year | Country,<br>Continent | Journal | Period | Age range | Trimester | Diagnosis method | Stone location (No.) | Stone size, mm<br>(mean/SD,range) |
| --- | --- | --- | --- | --- | --- | --- | --- | --- | --- |
| Ulvik[8] | 1995 | Norway,<br>Europe | Journal of Urology | September<br>1984-December<br>1994 | 27 (20-41) | 4-14 weeks in 3;<br>15-28 weeks in 9;<br>29-37 weeks in 12 | KUB 1 positive in 6;<br>US 3 positive in 21<br>(hydronephrosis 21 in 21) | Not mentioned | Not mentioned |
| Scarpa[9] | 1996 | Italy,<br>Europe | Journal of Urology | 3-years period | 24 (16-30) | 20-34 | US\symptoms\urinalysis | Not mentioned | Not mentioned |
| Parulkar[10] | 1998 | America,<br>North America | Journal of Urology | January 1984-November 1995 | 27 ( <18y 2; 18-20y 4; 20-30y<br>43; 30-40y 21) | First trimester in 3;<br>second trimester in 23;<br>third trimester in 44 | US 40 positive in 65;<br>IVP 5 positive in 5 | Not mentioned | US 0.7 (0.4-1.6);<br>IVP 0.55 (0.4-0.7) |
| Lemos[11] | 2002 | Brazil,<br>South America | International Braz J<br>Urol | Not mentioned | 28 (20-34) | 18 (12-34) | US 12 positive in 12;<br>ureteroscopy 13 positive in<br>14 | Proximal ureter in 1;<br>medium ureter in 4;<br>distal ureter in 12;<br>1 missed | 6 (4-12) |
| Rana[12] | 2009 | Pakistan,<br>Asia | Urology | 1997 - 2007 | 22 (18-27) | 20 (14-34)<br>First trimester in 1;<br>second trimester in 11;<br>third trimester in 7 | US in 11; KUB in 1 | Proximal ureter in<br>11;<br>distal ureter in 8; | 11 (8-18) |
| Elgamasy[13] | 2010 | Egypt,<br>Africa | BJU International | June 2003- June<br>2008 | 25.9 (18-38) | 25.9 (24-30) | US 12 positive in 15;<br>RU 14 positive in 15, | Proximal ureter in 2;<br>medium ureter in 2;<br>distal ureter in 10; | Not mentioned |
| Liu[14] | 2011 | China,<br>Asia | Journal of Huazhong<br>University of Science<br>and Technology-Medical Sciences | January 2004 -<br>December 2009 | 26.7 (18-37) | 23.45 (4-38) | US in 24 | 6 bilateral; 8 left; 10<br>right (surgery group) | Not mentioned |
| Polat[15] | 2011 | Turkey,<br>Asia | Urological Research | 2007-2009 | 25 (19-34) | 30 (23-35)<br>second trimester in 8;<br>third trimester in 8 | US in 11 | Proximal ureter in 5;<br>distal ureter in 6; | 9.45 (5-12) |
| Atar[16] | 2012 | Turkey,<br>Asia | International Journal<br>of Surgery | December<br>2010-July 2011 | 26 (19-40) | 24 (16-33) | US for 8, ureteroscopy for<br>all | Proximal ureter in 5;<br>medium ureter in 5;<br>distal ureter in 7;<br>no stone in 2 | 8 (5-19) |
| Bozkurt[17] | 2012 | Turkey,<br>Asia | Urological Research | April 2005-November 2010 | 27.8 (20-39) | 24 (15-34) | US 16 positive;<br>all 32 positive underwent<br>URS | Proximal ureter in 8;<br>medium ureter in 9;<br>distal ureter in 10;<br>no stone in 5 | 8 (5-19, in 16 US<br>positive cases) |
| Hoscan[18] | 2012 | Turkey,<br>Asia | Urology | 2001-2011 | 24 (17-37) | 26 (12-38) | URS 34 positive in 57 | Proximal ureter in 8;<br>medium ureter in 6;<br>distal ureter in 20 | 7 (4-13) |
| Johnson[19] | 2012 | America,<br>North America | Journal of Urology | Not mentioned | 27 | 24.7 | Low dose CT in 23;<br>US in 18;<br>MRI in 5 | Not mentioned | 7.8 (3-25) |
| Abdel[20] | 2013 | Egypt,<br>Africa | Urology Annals | April 2008-March 2011 | 23 (19-28) | 25 (16-35) | Clinical presentation and<br>US;<br>MRI in 3 | Proximal ureter in 2;<br>medium ureter in 5;<br>distal ureter in 10 | 17 (12-21) |
| Bozkurt[21] | 2013 | Turkey,<br>Asia | Urolithiasis | April 2005-Setemper 2011 | 27.41 ± 5.79 | 23.2 ± 4.6 (13-34) | Clinical presentation,<br>presence of microscopic<br>hematuria in urinalysis and<br>US | Proximal ureter in<br>13;<br>medium ureter in 13;<br>distal ureter in 15 | 9.78 ± 3.47 |
| Song[22] | 2013 | China, Asia | International Journal of Gynecology and Obstetrics | April 2001 - July 2012 | 27.2§<br>27.1¶ | 26.5§<br>26.3¶ | US 23 positive in 54;<br>MRI 25 positive in 31 | Proximal ureter in 10;<br>distal ureter in 44 | 13.14 (7-22) |
| Keshvari[23] | 2013 | Iran, Asia | Nephro-Urology Monthly | June 2003-April 2011 | 23 ± 2 (19-34) | 24 ± 3 (12-36)<br>First trimester in 2;<br>second trimester in 26;<br>third trimester in 16 | US in 44;<br>IVP in 2 | Proximal ureter in 2;<br>medium ureter in 10;<br>distal ureter in 36 | Not mentioned |
| Ngai[24] | 2013 | Iraq, Asia | Arab Journal of Urology | March 2008-March 2010 | 27.2 (18-38) | First trimester in 5;<br>second trimester in 15;<br>third trimester in 10 | US showed hydronephrosis in 30, stone in 12 | Not mentioned | Not mentioned |
| Adanur[25] | 2014 | Turkey, Asia | Archivio Italiano di Urologia e Andrologia | January 2005-December 2012 | 25.4 (18-41) | 24.8(7-33) | US in 6 ;<br>ureteroscopy for all | Proximal ureter in 6;<br>medium ureter in 5;<br>distal ureter in 8 | 9.2 (6-13) in 6 with US |
| Georgescu[26] | 2014 | Romania, Europe | Chirurgia | January 2006-January 2012 | 27.2 (20-37) | First trimester in 6;<br>second trimester in 32;<br>third trimester in 16 | US stone 18 positive in 54 | Proximal ureter in 11;<br>medium ureter in 8;<br>distal ureter in 14<br>Not mentioned | 8 (4-16) |
| Teleb[27] | 2014 | Egypt, Africa | Arab Journal of Urology | October 2006-December 2013 | 26.6 (SD 4.65)§<br>25.5 (SD 4.26)¶ | 24.1 (SD 5.44)§<br>25.7 (SD 4.95)¶ | US 31 positive in 43 | Middle ureter in 9§;<br>distal ureter in 13§<br>Middle ureter in 7¶;<br>distal ureter in 14¶ | Not mentioned |
| Wang[28] | 2014 | China, Asia | Urology | February 2006-Setemper 2012 | 26 (17-39) | 29(17-39)<br>First trimester in 2;<br>second trimester in 36;<br>third trimester in 49 | US in 79, MRI in 8, | Left side in 48, Right side in 39 | 8 (5-19) |
| Fathelbab[29] | 2016 | Egypt, Africa | African Journal of Urology | April 2006-October 2013 | 23 (19-37) | First trimester in 4;<br>second trimester in 23;<br>third trimester in 14 | Diagnostic ureteroscopy<br>36 positive in 41 | Proximal ureter in 7;<br>distal ureter in 29 | 8.9 (5-16) |
| Zhang[30] | 2016 | China, Asia | PLoS ONE | March 2009-Setemper 2014 | 25.5±4.6 (16–41) | 9-36 | US and diagnostic ureteroscopy positive in 86 (only ureteroscopy in 24), negative in 31 | Not mentioned | 8.2 ± 0.6 |
| Abedi[31] | 2017 | Iran, Asia | Journal of Lasers in Medical Sciences | January 2007-June 2016 | 29.3 | 27.3 (13-31)<br>First trimester in 9;<br>second trimester in 24;<br>third trimester in 12 | Clinical manifestations, urinalysis and US | Poximal ureter in 5;<br>distal ureter in 40 | 7.84 (5-9mm) |
| Tan[32] | 2018 | China, Asia | European Journal of Obstetrics and Gynecology and Reproductive Biology | January 2005-June 2015 | 26.7 ± 8.9§<br>27.4 ± 10.2¶ | 27.5 ± 11.2§<br>25.9 ± 9.7¶ | US | Proximal ureter in 10;<br>medium ureter in 12;<br>distal ureter in 31 | Not mentioned |

§ means received internal ureteral stent only; ¶ means received ureteroscopy operation.

### Subgroup analysis and meta-analysis

There were 2 studies involving D-J stent insertion only[10, 24], 19 studies involving URS operation[8, 9, 11-21, 23, 25, 26, 29-31], and 4 involving both[22, 27, 28, 32]. A total of 131 cases undergoing internal ureteral stent only and 789 cases undergoing URS operation. Common results were showed in tables and occurrence rates (ORs) were calculated and compared by meta-analysis.

Detailed data of internal ureteral stent therapy was showed in **Table 2**. The most commonly used anesthesia was local anesthesia. The pooled operation success rate was 97% [**Figure 2**, 95% CI, 0.94-1.01]. Only one related study mentioned the SFR was about 25% [**Figure 3**, 95% CI, 0.04-0.46], which reported as an accident situation. The pooled ORs of normal fertility outcome was 99% [**Figure 4**, 95% CI, 0.99-1.01] and the pooled ORs of adverse pregnant outcome (premature and abortion) was less than 1% [**Figure 5**, 95% CI, 0-0.02]. The pooled ORs of overall complications was about 45% [**Figure 6**, 95% CI,0.19-0.70], but the pooled ORs of serious complications (Clavien-Dindo III-V) was less than 1% [**Figure 7**, 95% CI,0-0].

**Table 2.** Summary of details for D-J stent therapy group.

| First author | Year | Anesthetic method | No. of operations<br>(success rate) | SFR, % | Fertility outcome | Complications | Complications<br>(classified) | Follow-up pattern |
| --- | --- | --- | --- | --- | --- | --- | --- | --- |
| Parulkar[10] | 1998 | Local anesthesia | 15 (100%) | \ | Not mentioned | Stent slipping into bladder in 1, then repeaced;<br>5F stent blocked in 2,then replace to 7F;<br>softer stent was needed in 1;<br>calcified stent in 1 | Clavien-Dindo III in 5 | Not mentioned |
| Song[22] | 2013 | Local anaesthesia with<br>lidocaine gel | 17, 12 success<br>(70.6%) | 25 (3 passed stone<br>spontaneously of<br>12) | 16 delivered at term;<br>preterm labor in 1 | Stent-induced bladder irritation in 6, retained;<br>encrusted stent problem in 4;<br>passed a double-J stent in 1 | Clavien-Dindo I in 6;<br>Clavien-Dindo III in 5; | Not mentioned |
| Ngai[24] | 2013 | Local anaesthesia | 30 (100%) | \ | Not mentioned | Stent encrustation in 3;<br>stent migration in 3;<br>stent-related bladder irritation in 3;<br>gross hematuria in 2 | Clavien-Dindo I in 5;<br>Clavien-Dindo III in 6 | Renal function tests and US was<br>arranged weekly in the first<br>month, then monthly throughout<br>pregnancy |
| Teleb[27] | 2014 | Spinal anaesthesia in 18,<br>topical lidocaine anaesthesia<br>with sedo-analgesia in 4 | 22 (100%) | \ | All 22 delivered at<br>term | Urinary tract infection in 4;<br>irritative LUTS in 13 | Clavien-Dindo I in 13;<br>Clavien-Dindo II in 4 | US and urinalysis every 4 weeks |
| Wang[28] | 2014 | Epidural anesthesia | 17 (100%) | \ | All 17 delivered at<br>term | Urinary tract infection in 4;<br>stent-related bladder irritation in 12;<br>hemauria in 7 | Clavien-Dindo I in 19;<br>Clavien-Dindo II in 4 | Obstetric care; clinical<br>assessment, ultrasound<br>examination and urine culture. |
| Tan[32] | 2018 | Local anesthesia | 30, 25 success<br>(83.3%) | \ | Not mentioned | Bladder irritation in 2;<br>D-J stent drop in 1;<br>hard removal of D-J stent in 1 | Clavien-Dindo I in 3;<br>Clavien-Dindo III in 1 | Not mentioned |

SFR: stone-free rate.

**Figure 2.**
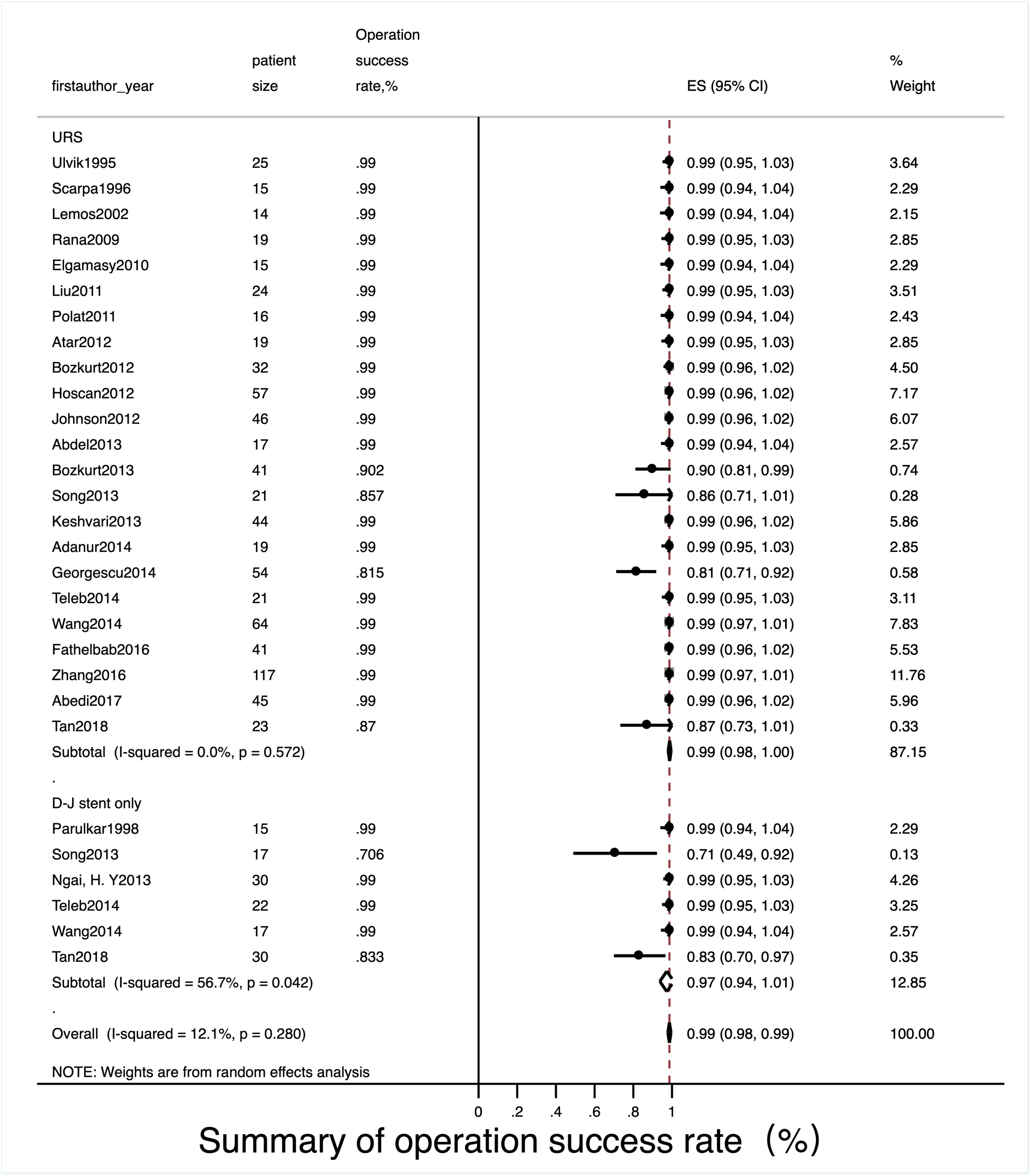
Meta-analysis about operation success rate in D-J stent therapy group and URS group.

**Figure 3.**
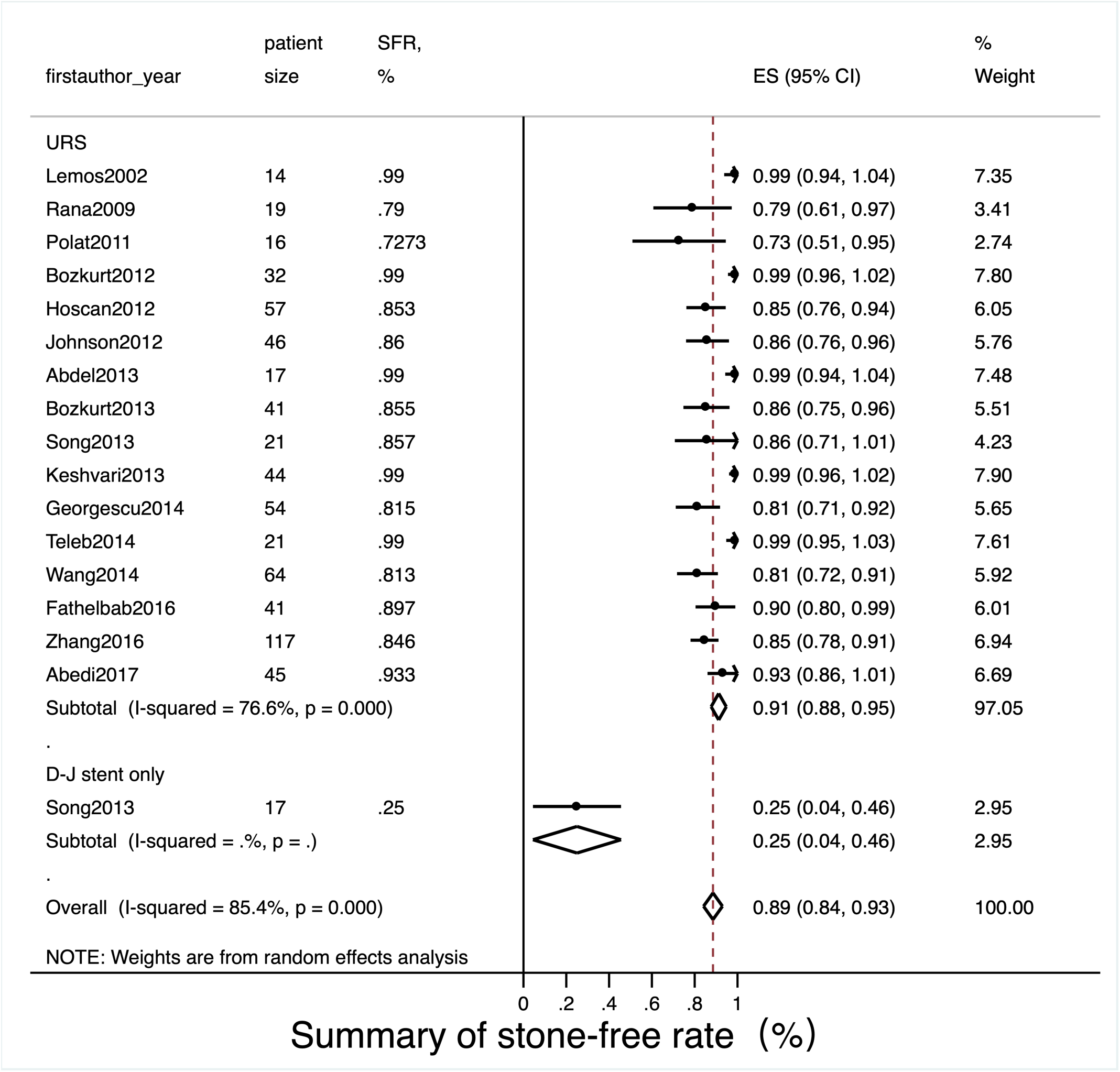
Meta-analysis about stone free rate in D-J stent therapy group and URS group.

**Figure 4.**
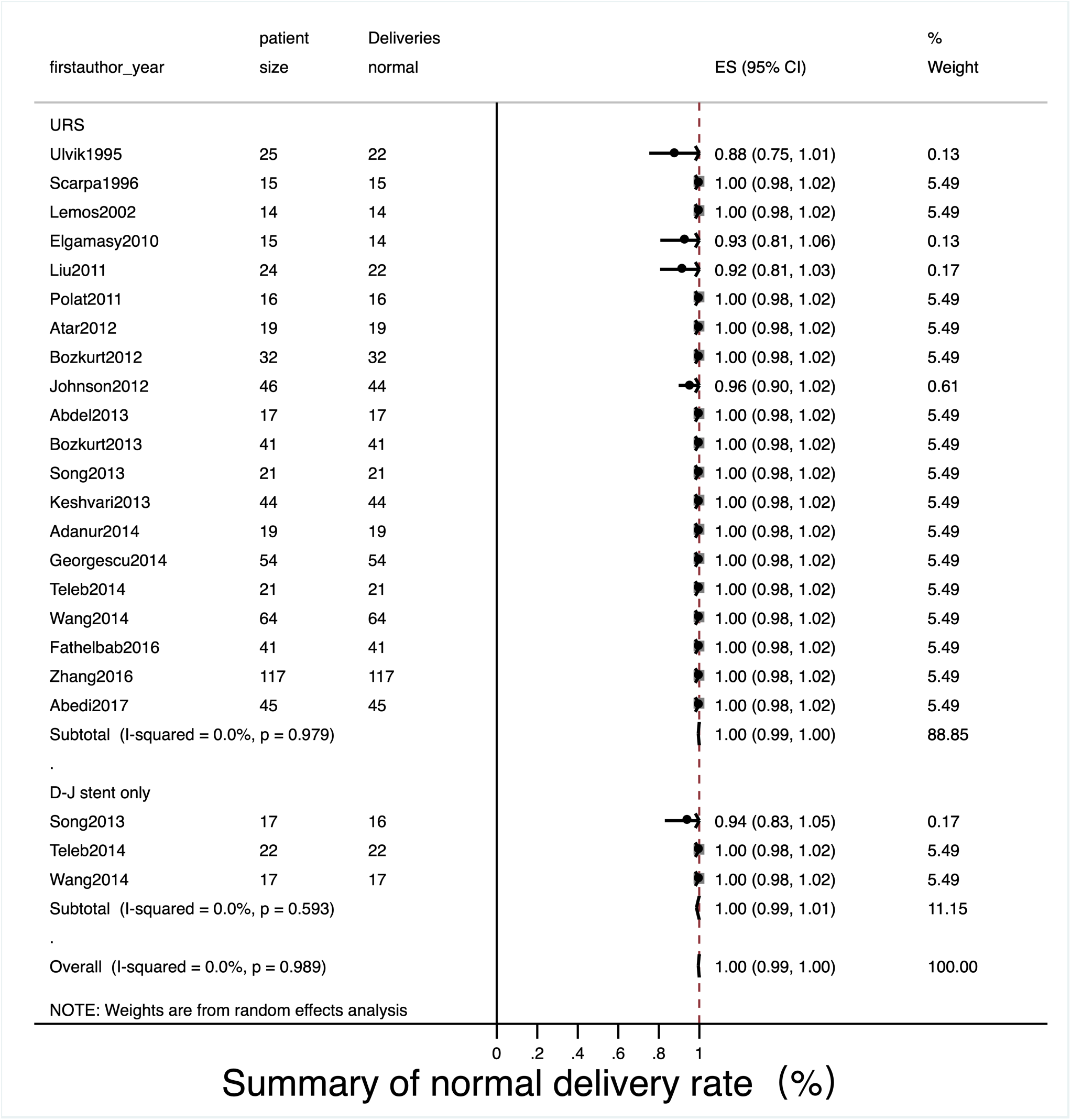
Meta-analysis about normal fertility outcome in D-J stent therapy group and URS group.

**Figure 5.**
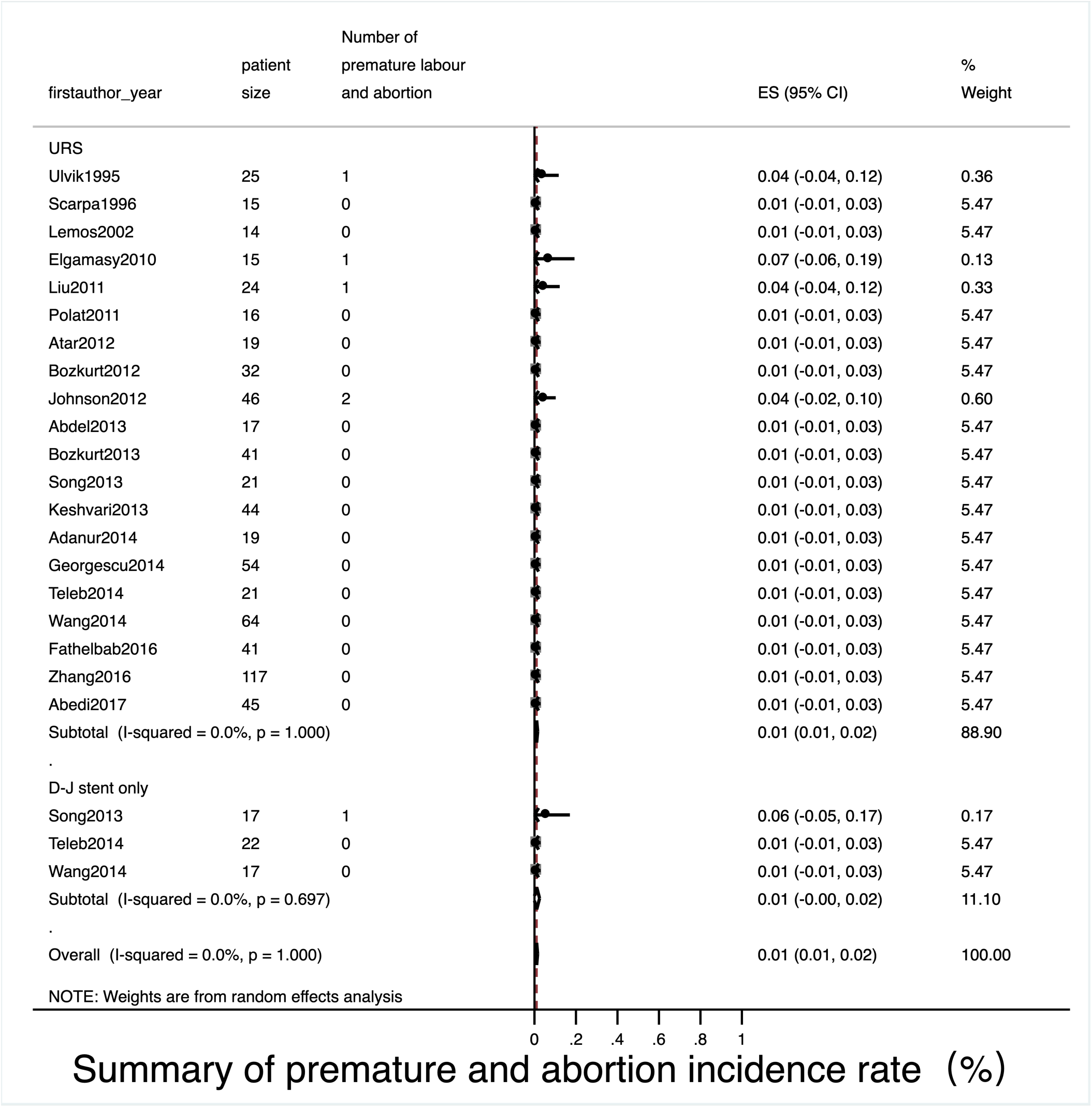
Meta-analysis about adverse pregnant outcome (premature and abortion) in D-J stent therapy group and URS group.

**Figure 6.**
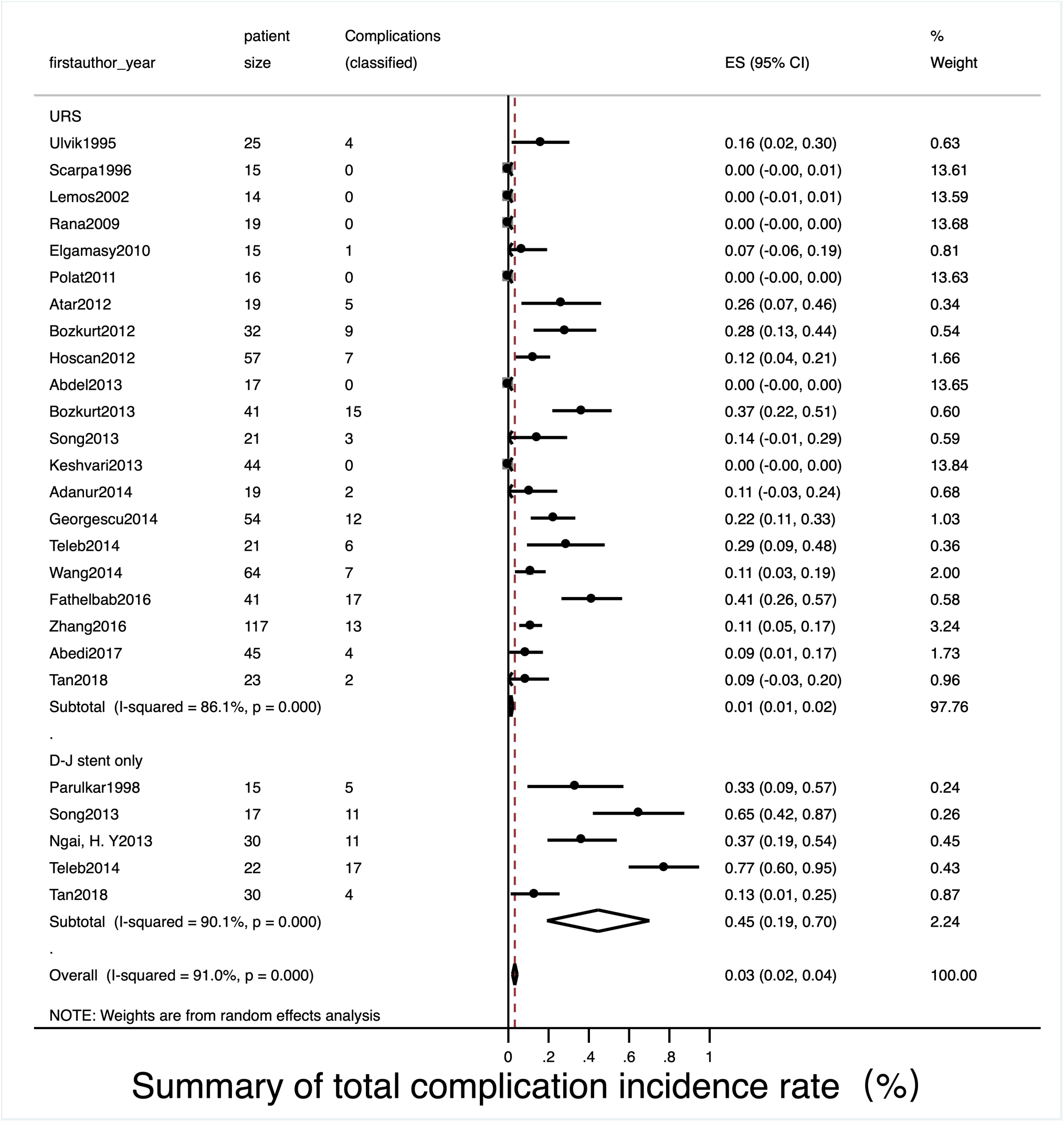
Meta-analysis about overall complications in D-J stent therapy group and URS group.

**Figure 7.**
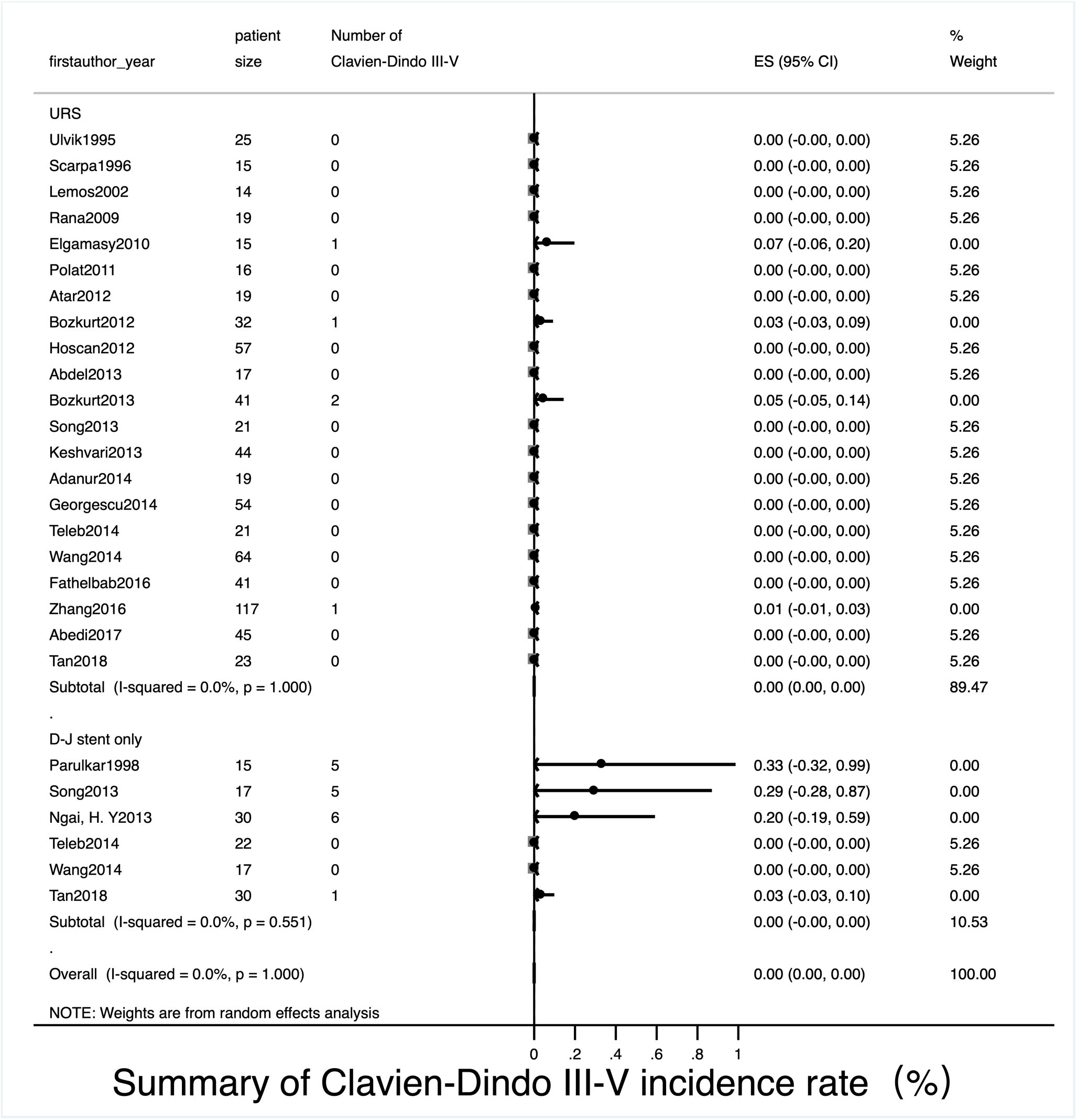
Meta-analysis about Clavien-Dindo III-V complications in D-J stent therapy group and URS group.

Detailed data of URS therapy was showed in **Table 3**. General anesthesia and spinal anesthesia were widely used in this situation. The pooled operation success rate was 99% [**Figure 2**, 95% CI, 0.98-1]. The SFR was about 91% in all [**Figure 3**, 95% CI, 0.88-0.95]. The pooled ORs of normal fertility outcome was 99% [**Figure 4**, 95% CI, 0.99-1], the pooled ORs of adverse pregnant outcome was less than 1% [**Figure 5**, 95% CI, 0.01-0.02]. The pooled ORs of overall complications was about 1% [**Figure 6**, 95% CI, 0.01-0.02], and the pooled ORs of serious complications (Clavien-Dindo III-V) was less than 1% [**Figure 7**, 95% CI,0-0].

**Table 3.**
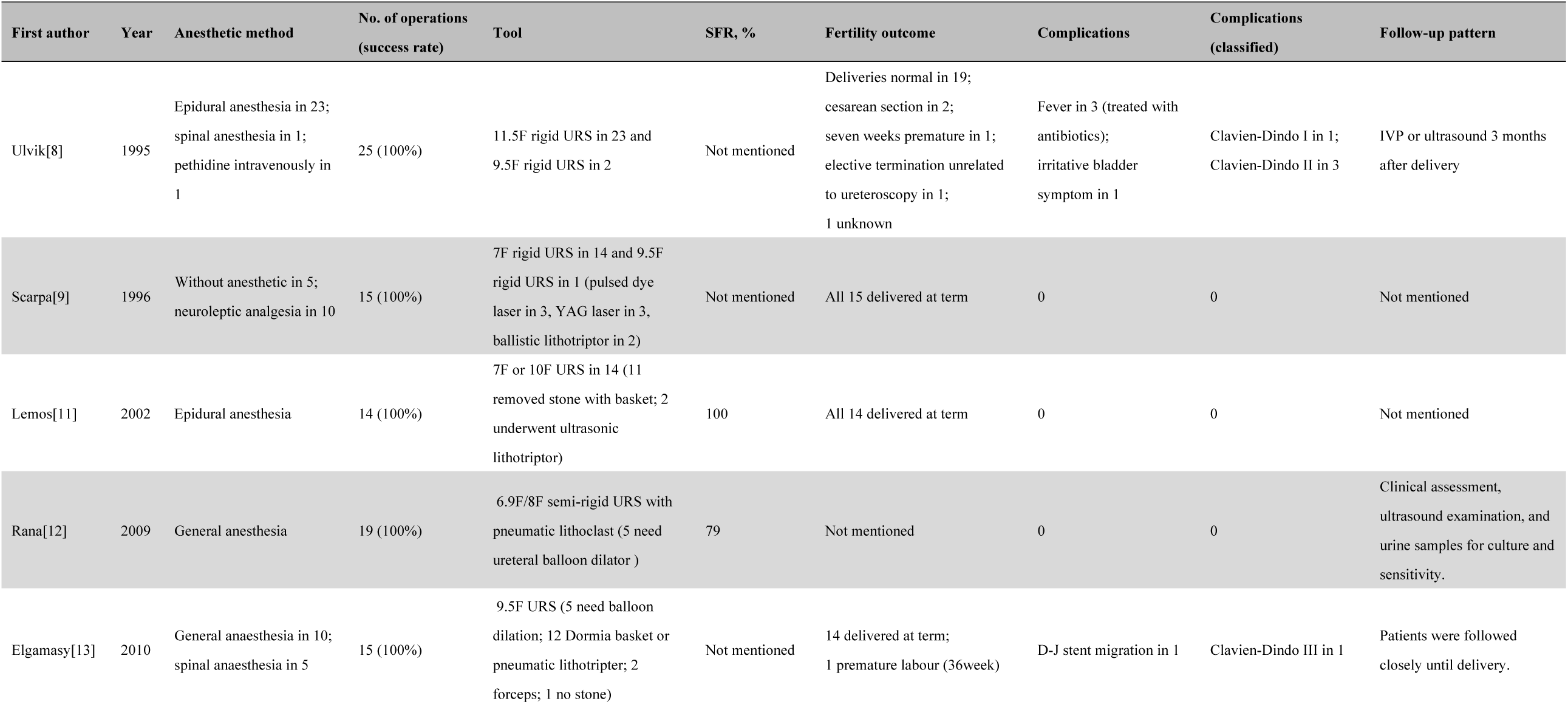
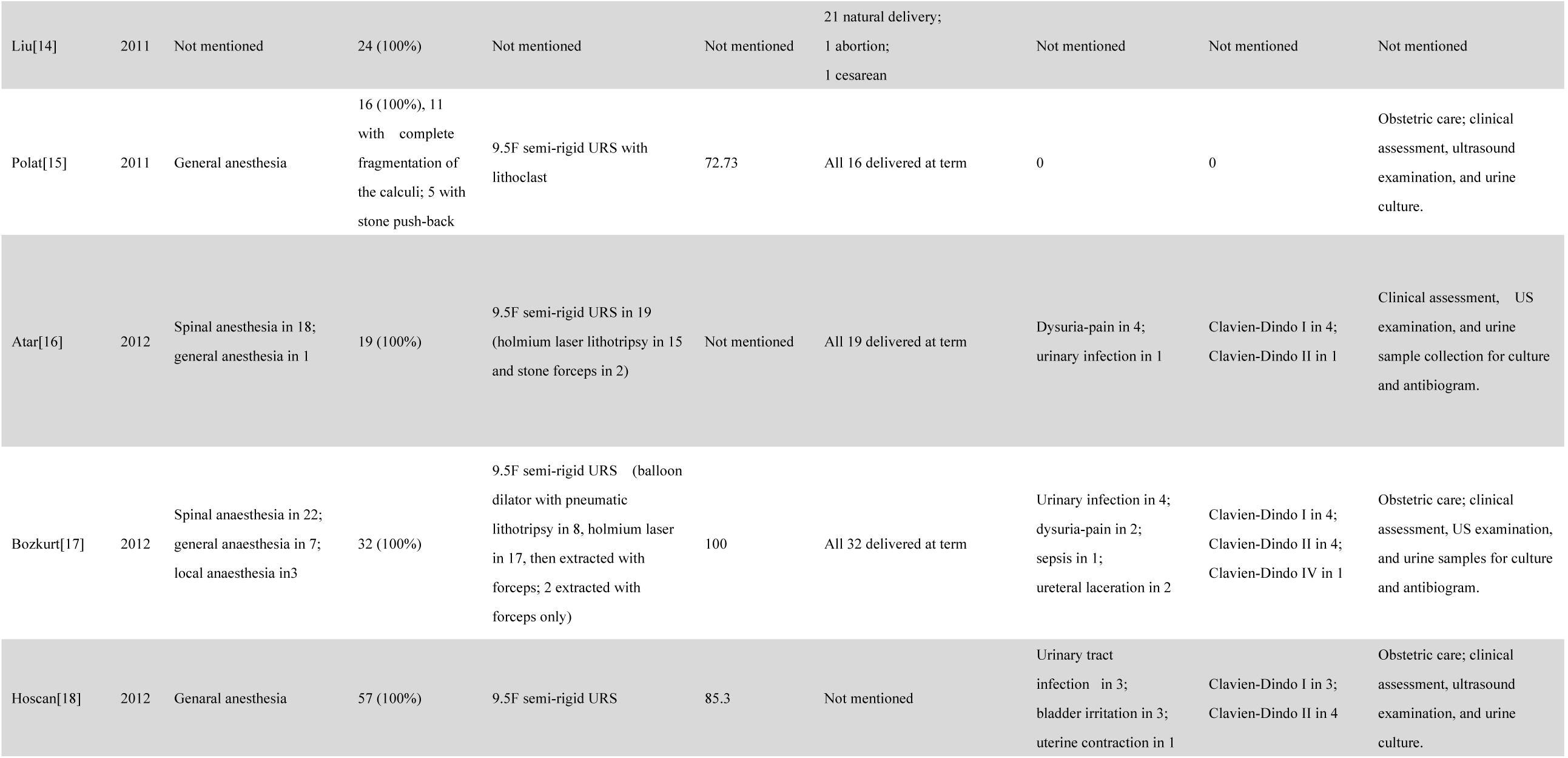
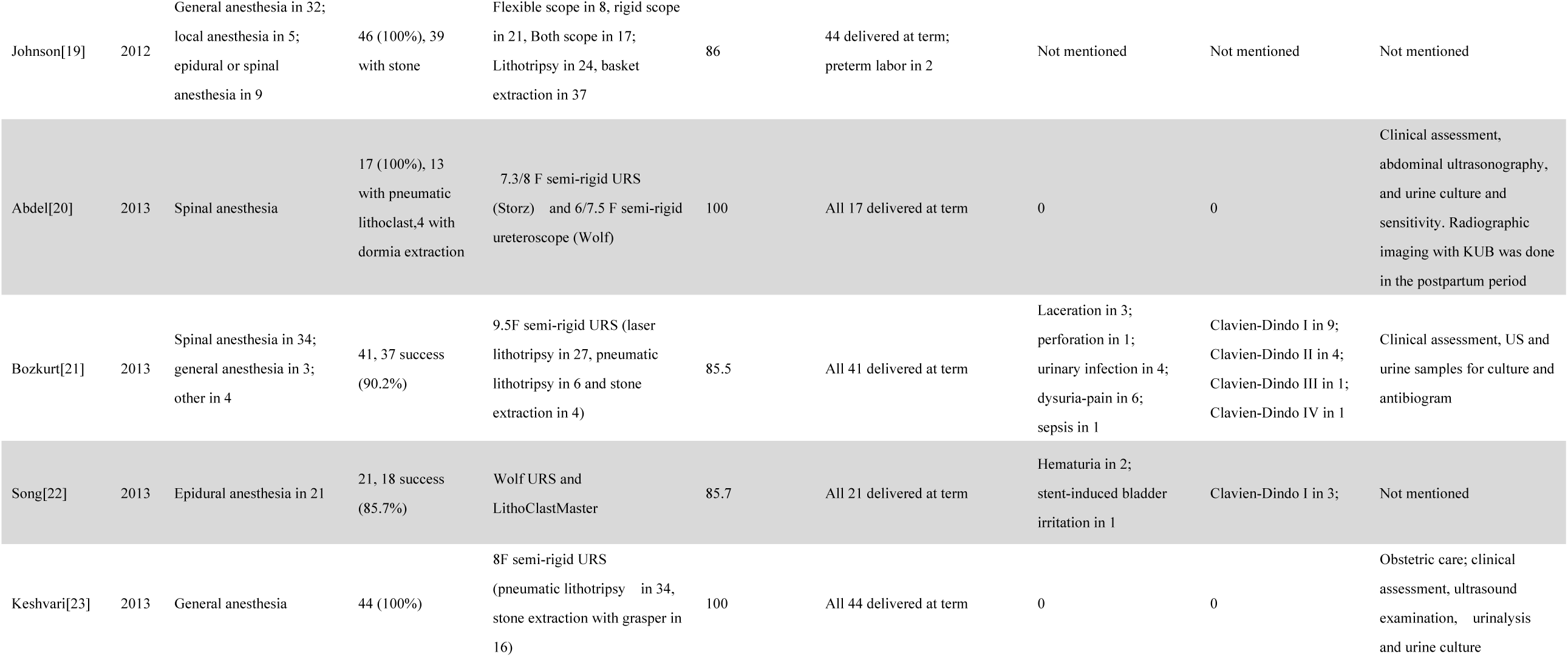
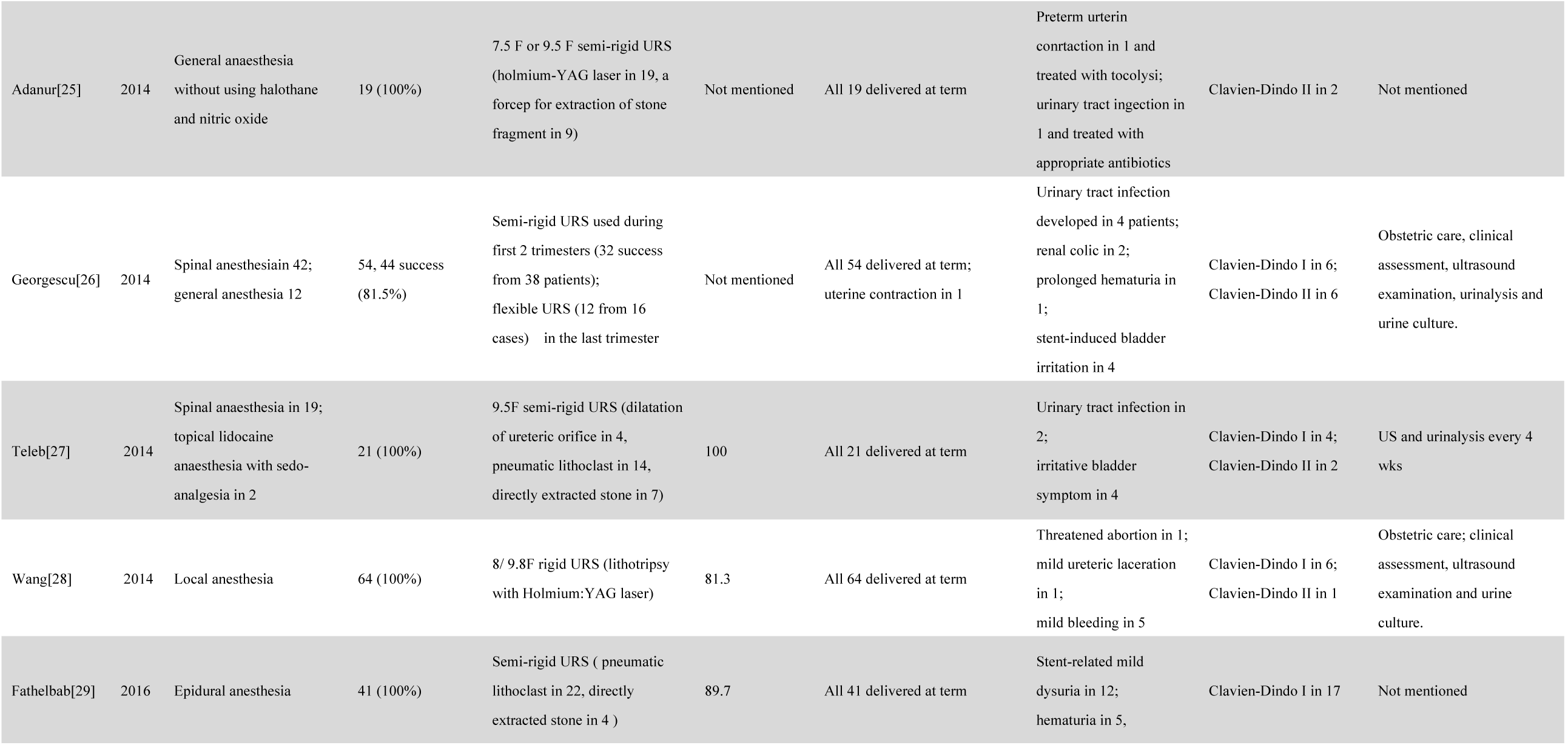
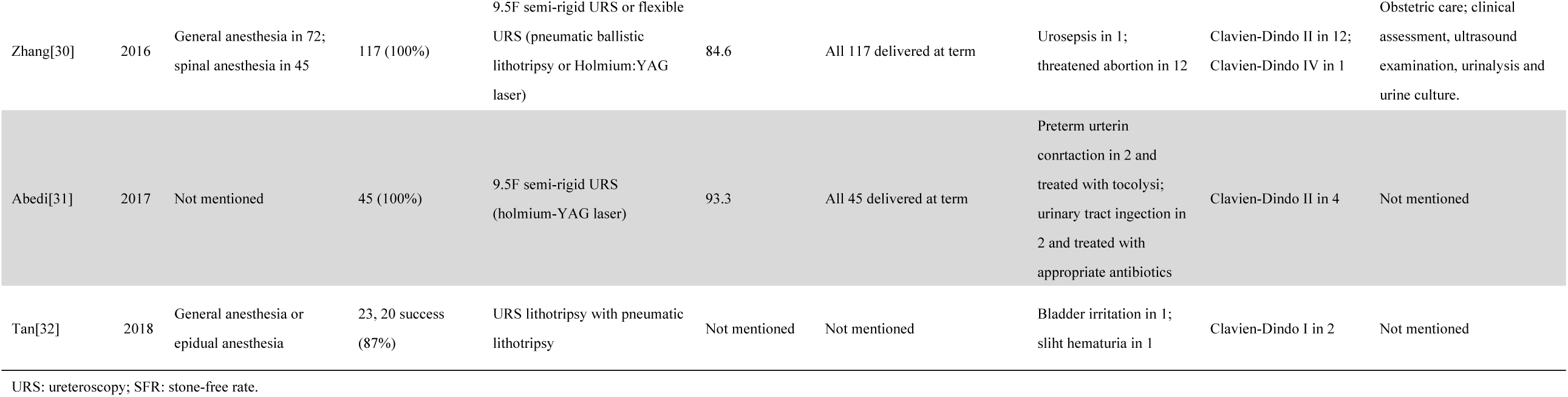
Summary of details for URS group.

Meta-analysis results indicated that there was no evidence of statistical heterogeneity between two treatments on operation success rate (**Figure 2**, *I*^2^=12.1%, P=0.280), normal fertility outcome (**Figure 4**, *I*^2^=0.0%, P=0.989) and adverse pregnant outcome (**Figure 5**, *I*^2^=0.0%, P=1.000). However, overall complications of internal ureteral stent therapy was more common than that in URS operation group (**Figure 6**, *I*^2^=91.0%, P < 0.001). We also analyzed pooled ORs of serious complications in two treatments (**Figure 7**). There was no evidence of significant statistical heterogeneity among studies (*I*^2^=0.0%, *P*=1.000).

### Qualitative assessment and publication bias

The NOS tool was used to conduct a qualitative assessment of the selected studies to review the quality of the studies and detect possible bias (**Table 4** and **Table 5**). Of the 25 studies, 8 were at low risk of bias (7-9 stars). 16 studies were at medium risk (4-6 stars) mainly due to bias from representativeness of case or controls, control definition and comparability. 1 study was high risk (3 stars) mainly due to bad representativeness, lack of control and unclear control exposure. The funnel plot showed certain publication bias in the studies included in the meta-analysis (Begg’s test with P<0.001) (Figure S1).

**Table 4.** Newcastle-Ottawa Scale review for cohort studies from systematic review

| Study | Country | Selection |  |  |  | Comparability |  | Outcome |  |  | Total |
| --- | --- | --- | --- | --- | --- | --- | --- | --- | --- | --- | --- |
|  |  | S1 | S2 | S3 | S4 | C1 | C2 | O1 | O2 | O3 |  |
| Liu et al. [14] | China | ★ | ★ | ★ | ★ |  |  | ★ | ★ | ★ | 7 |
| Bozkurt et al.[17] | Turkey | ★ | ★ | ★ | ★ |  |  | ★ | ★ | ★ | 7 |
| Teleb et al.[27] | Egypt | ★ | ★ | ★ | ★ |  |  | ★ | ★ | ★ | 7 |

Guidelines for review
*Selection:*
S1, Representativeness of the exposed cohort: ★a) representative of the community (e.g. community-based colorectal cancer-screening programme or registry) or (single hospital or clinic); b) selected group of people (e.g. nurses, volunteers); d) no description of the derivation of the cohort
S2, Selection of the non-exposed cohort: ★a) drawn from the same community as the exposed cohort; b) drawn from a different source; c) no description of the derivation of the non-exposed cohort
S3, Ascertainment of exposure: ★ a) secure record (eg medical records); ★b) structured interview; c) written self-report; d) no description
S4, Demonstration that outcome of interest was not present at start of study: ★ a)yes; b) no
*Comparability:*
C1, ★ Study controls for one most important factor;
C2, ★ Study controls for any additional factors (1> additional factors)

**Table 5.**
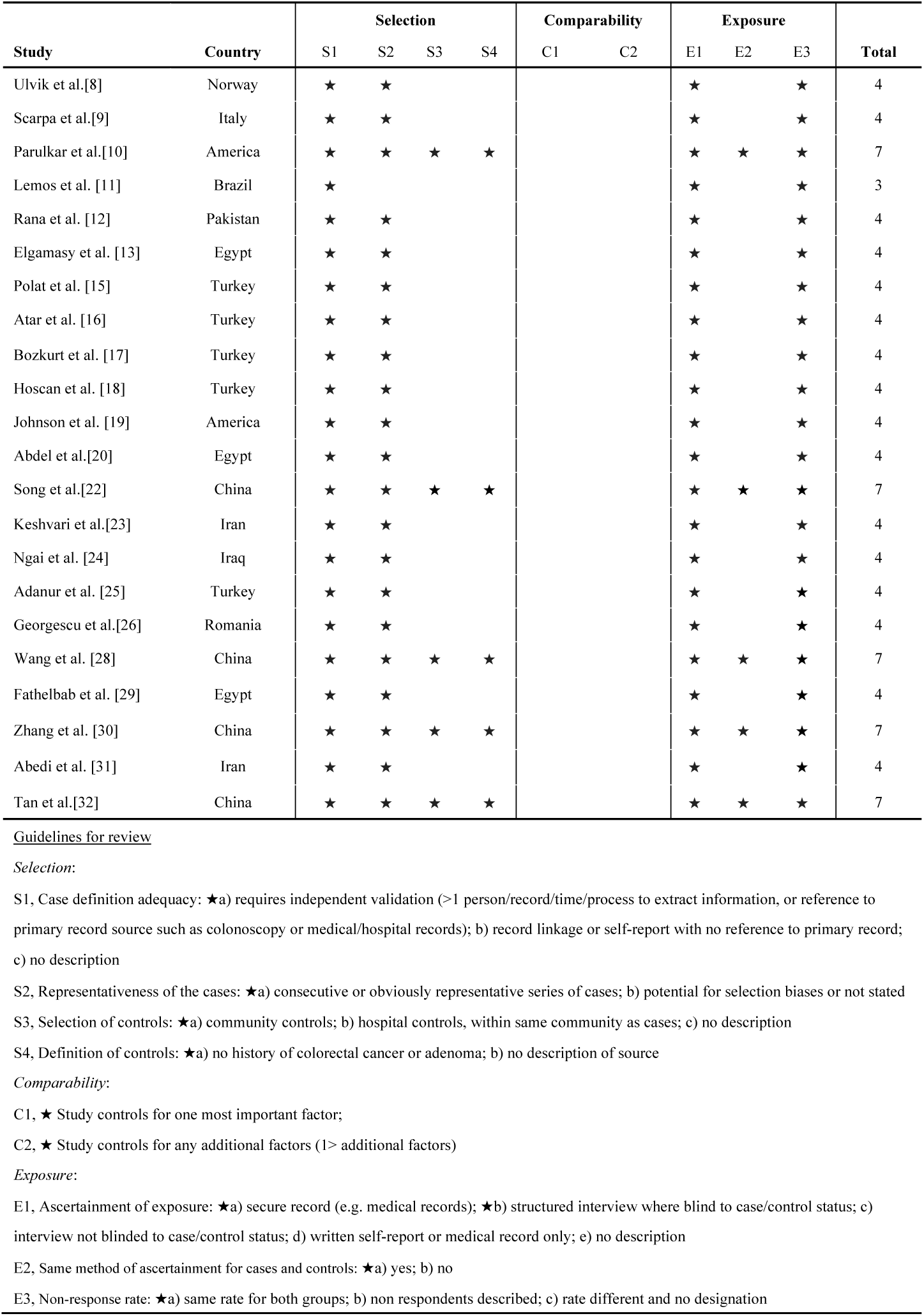
Newcastle-Ottawa Scale review for case-control and cross-sectional studies from systematic review

| Study | Country | Selection |  |  |  | Comparability |  | Exposure |  |  | Total |
| --- | --- | --- | --- | --- | --- | --- | --- | --- | --- | --- | --- |
|  |  | S1 | S2 | S3 | S4 | C1 | C2 | E1 | E2 | E3 |  |
| Ulvik et al.[8] | Norway | ★ | ★ |  |  |  |  | ★ |  | ★ | 4 |
| Scarpa et al.[9] | Italy | ★ | ★ |  |  |  |  | ★ |  | ★ | 4 |
| Parulkar et al.[10] | America | ★ | ★ | ★ | ★ |  |  | ★ | ★ | ★ | 7 |
| Lemos et al. [11] | Brazil | ★ |  |  |  |  |  | ★ |  | ★ | 3 |
| Rana et al. [12] | Pakistan | ★ | ★ |  |  |  |  | ★ |  | ★ | 4 |
| Elgamasy et al. [13] | Egypt | ★ | ★ |  |  |  |  | ★ |  | ★ | 4 |
| Polat et al. [15] | Turkey | ★ | ★ |  |  |  |  | ★ |  | ★ | 4 |
| Atar et al. [16] | Turkey | ★ | ★ |  |  |  |  | ★ |  | ★ | 4 |
| Bozkurt et al. [17] | Turkey | ★ | ★ |  |  |  |  | ★ |  | ★ | 4 |
| Hoscan et al. [18] | Turkey | ★ | ★ |  |  |  |  | ★ |  | ★ | 4 |
| Johnson et al. [19] | America | ★ | ★ |  |  |  |  | ★ |  | ★ | 4 |
| Abdel et al.[20] | Egypt | ★ | ★ |  |  |  |  | ★ |  | ★ | 4 |
| Song et al.[22] | China | ★ | ★ | ★ | ★ |  |  | ★ | ★ | ★ | 7 |
| Keshvari et al.[23] | Iran | ★ | ★ |  |  |  |  | ★ |  | ★ | 4 |
| Ngai et al. [24] | Iraq | ★ | ★ |  |  |  |  | ★ |  | ★ | 4 |
| Adanur et al. [25] | Turkey | ★ | ★ |  |  |  |  | ★ |  | ★ | 4 |
| Georgescu et al.[26] | Romania | ★ | ★ |  |  |  |  | ★ |  | ★ | 4 |
| Wang et al. [28] | China | ★ | ★ | ★ | ★ |  |  | ★ | ★ | ★ | 7 |
| Fathelbab et al. [29] | Egypt | ★ | ★ |  |  |  |  | ★ |  | ★ | 4 |
| Zhang et al. [30] | China | ★ | ★ | ★ | ★ |  |  | ★ | ★ | ★ | 7 |
| Abedi et al. [31] | Iran | ★ | ★ |  |  |  |  | ★ |  | ★ | 4 |
| Tan et al.[32] | China | ★ | ★ | ★ | ★ |  |  | ★ | ★ | ★ | 7 |

Guidelines for review*Selection:*
S1, Case definition adequacy: ★a) requires independent validation (>1 person/record/time/process to extract information, or reference to primary record source such as colonoscopy or medical/hospital records); b) record linkage or self-report with no reference to primary record; c) no description
S2, Representativeness of the cases: ★a) consecutive or obviously representative series of cases; b) potential for selection biases or not stated
S3, Selection of controls: ★a) community controls; b) hospital controls, within same community as cases; c) no description
S4, Definition of controls: ★a) no history of colorectal cancer or adenoma; b) no description of source
*Comparability:*
C1, ★ Study controls for one most important factor;
C2, ★ Study controls for any additional factors (1> additional factors)
*Exposure:*

## DISCUSSION

From the best of our knowledge, that this study is the first systematic review to investigate and compare between the outcomes of the ureteroscopy and serial D-J stenting therapy in pregnancy with urolithiasis. To determine the efficacy and safety of two treatments, we analysed the relative information as much detail as possible. This meta⍰Ianalysis contained 25 studies with total 920 cases of urolithiasis during pregnancy. This meta⍰Ianalysis contains studies selected from several countries as stated above. As showed in **Table 1**, most studies come from Asia continent (15 studies), followed by Africa continent (4 studies), Europe continent (3 studies) and America continent (including North and South America, 3 studies). So this review could represent human races of different skin colors.

Urolithiasis in pregnancy is the most common cause of non-obstetric reason for hospital admission, 80–90% of which are diagnosed in the 2nd or 3rd trimester of their pregnancy when the disease becomes symptomatic[33-36]. As a majority of calculi could be passed with treatment of intravenous fluids and analgesia, the first-line treatment of urolithiasis in pregnancy is conservative management. This is recommended by both the latest European Association of Urology (EAU) and the American Urological Association (AUA). However, if complications develop and may even affect fetal safe, or the patient does not feel adequate relief, more aggressive treatment should be considered. Shock wave lithotripsy is absolutely contraindicated in pregnancy because of potential fetal death[37]. Percutaneous nephrostomy (PCN) drainage is also not an appropriate choice as it raises risk of septic complications and imposes additional burden of an external drain[38]. The common utilization of prone position and fluoroscopy limited PCN in pregnancy as well[39]. Therefore, internal ureteral stent and URS are the most common treatments in clinic for the pregnant patient.

After failed in initial conservative treatment, insertion of D-J stent might be a safe choice. Serial stenting for pregnancy with urolithiasis was common used in clinic but there were not many related studies. After scanning articles in the past 30 years, only 6 related articles were included in this meta-analysis[10, 22, 24, 27, 28, 32]. Historically, serial stenting was considered as the gold standard surgical treatment for pregnancy with urolithiasis as it was less invasive and could be performed under local anesthesia[40]. This amount of anesthetic drugs and fewer surgical traumas was safer for the fetus[24]. And its effect of relieving obstruction and pain, maintenance of pregnancy was proved as this meta-analysis suggested. But there were still some negative opinions. On the one hand, serial stenting may be poorly tolerated by some pregnant women as it caused pain and reducing quality of life. On the other hand, insertion of D-J stent was a temporary measure, the D-J stents need a regularly replacement. And due to the increased concentration of calcium and urate in urine during pregnancy, which cause more prone to encrustation, these invasive operations need more frequency[20, 41]. With the increase of invasive operations, complications such as UTI, stent migration were increased[27, 32, 42], and the cost raised as well[39]. Actually, our meta-analysis had demonstrated the pooled ORs of complications after serial stenting was about 45%. However, the pooled ORs of serious complications (Clavien-Dindo III-V) after serial stenting was less than 1%. And there is no evidence that serial stenting treatment was harmful for pregnancy as the pooled ORs of adverse pregnant outcome was less than 1%. Internal ureteral stent was proved to be safe for pregnant woman and fetus in all.

Not the same as internal ureteral stent operation, URS for treating urolithiasis in pregnancy were studied by many urologists, as 23 papers were included in this meta-analysis as mentioned above[8, 9, 11-23, 25-32]. It is common that anesthesia methods were including general anesthesia and spinal anesthesia after scanning the papers included. Although there were risks in anesthesia and surgery, development in technology provided a guarantee for perioperative safety. After systematic analysis, we calculated that the pooled ORs of complications was about 1% and the pooled ORs of normal fertility outcome was 99%. Another advantage of URS was the high SFR which arrived 91%. High stone clearance rates and low complication rates made URS be recommended by the 2020 EAU guideline.

In the latest 2020 EAU guideline, URS looks like a better selection for pregnancy with urolithiasis compared with internal ureteral stent, and stent insertion therapy is only mentioned for symptomatic moderate-to-severe hydronephrosis during pregnancy. It looks like ureteral stent insertion is not a proper treatment for pregnant women with urolithiasis. But we need to under that a successful URS operation must base on detailed preoperative preparation and stringent obstetric care. At emergency condition, or in a backward obstetric care environment, internal ureteral stent may be better choice as it is also safe and effective in all. And it could gain time for URS later. Urologist and obstetrician should work together to ensure the safety of pregnancy and fetus.

There were several inherent limitations in this meta⍰Ianalysis. First, most of the included studies were retrospective study. This might cause inevitable methodological defects in these studies, including data bias, insufficient baseline comparisons, and insufficient data collection. Urolithiasis during pregnancy is not a rare disease, but for urologists, it is not easy to handle both urolithiasis condition and obstetric care; and after failed in initial conservative treatment, it may be considered as an emergency to handle rapidly. Thus well-designed RCTs were difficult to accomplish. Secondly, performance bias should also be considered. Although various centres have performed similar operations, the medical equipment and medical teams were different. Surgery is a complex process; these differences may also lead to different outcome. What’s more, there was unavoidable bias when the data were pooled. Therefore, further well-designed, prospective studies are required, and those studies should take into account selection bias, performance bias and the issue of confounding. Finally, funnel plot showed certain publication bias in included articles, but considering the number of included article was small, we reserved all studies. Despite these limitations, this updated meta⍰Ianalysis provides an important clinical reference for the urolithiasis during pregnancy.

## CONCLUSION

Both ureteroscopy operation and internal ureteral stent were usually used for handing pregnancy with urolithiasis. Two treatments had less side effective on fertility outcome, but internal ureteral stent may cause more complications. Evidence suggests that URS therapy may have a greater advantage for pregnancy with urolithiasis when the conditions permit. As it is proved safe and effective, internal ureteral stent could be considered at emergency condition or preoperative preparations was lack.

## Data Availability

All data relevant to the study are included in the article or uploaded as supplementary information.

## Figure legends/captions

**Figure S1**. Funnel Plot for Publication Bias.

**Table S1**. Search strategy and results.

**Table S2**. Complications and their Clavien-Dindo Classification.

